# A multimodal biomarker predicts dissemination of bronchial carcinoid

**DOI:** 10.1101/2021.05.17.21257308

**Authors:** E.M.B.P. Reuling, D.D. Naves, E. Thunnissen, P.C. Kortman, M.A.M.B. Broeckaert, P.W. Plaisier, C. Dickhoff, J.M.A. Daniels, T. Radonic

**Affiliations:** Department of Surgery, De Boelelaan 1117, 1081 HV Amsterdam, the Netherlands; Department of Cardiothoracic Surgery, De Boelelaan 1117, 1081 HV Amsterdam, the Netherlands; Department of Pathology, De Boelelaan 1117, 1081 HV Amsterdam, the Netherlands; Department of Pulmonary Diseases, Amsterdam University Medical Center, VU University Amsterdam, De Boelelaan 1117, 1081 HV Amsterdam, the Netherlands; Cancer Centre Amsterdam; De Boelelaan 1117, 1081 HV Amsterdam, the Netherlands; Department of Surgery, Albert Schweitzer Hospital, Albert Schweitzerplaats 25, 3318 AT Dordrecht, the Netherlands

**Keywords:** Typical carcinoid, atypical carcinoid, lymph node metastases, distant metastases, OTP, CD44, P16, Rb, Ki-67, mitotic count, prognosis

## Abstract

**Introduction:** The extensive loss of lung parenchyma is a drawback of anatomical resection in bronchial carcinoids. Endobronchial therapy (EBT) has emerged as a safe and effective minimally invasive tissue sparing alternative for small intraluminal tumors. Currently, therapeutic decision making in patients with bronchial carcinoid is mainly based on tumor morphology and patient characteristics. The availability of more accurate biomarkers might help clinicians in selecting low-risk tumors for EBT. Therefore, we investigated radiological (tumor diameter), morphometric (mitotic index) and immunohistochemical (OTP, CD44, Ki-67, Rb and P16) markers as predictors of dissemination.

**Material and methods:** Patients referred to Amsterdam University Medical Centers with available histology were included. Clinical and morphological characteristics relevant for classification such as tumor diameter, mitotic count (MAI) and prognostic immunohistochemical markers as Ki-67, P16, Rb, Orthopedia homebox (OTP) and CD44 were analyzed.

**Results:** In a cohort of 171 patients, the vast majority were curatively treated with either EBT (*n*=61, 36%) or surgery (*n*=103, 60%). Seven (4%) patients presented with distant metastases at diagnosis. TC was diagnosed in 112 (65%) and AC in 59 (35%) patients. Nine (15%) patients treated with EBT had a local recurrence of disease during follow up and none developed lymph node or distant metastasis. Of all surgically treated patients, 13 (13%) had level 1 or 2 lymph node metastases. Additional 13 (13%) patients developed distant metastases, 11 (85%) were AC and 2 (15%) TC. Patients with tumor stage IA (tumor diameter ≤1cm) irrespective of tumor classification or immunohistochemical results did not develop distant metastases. Patients with typical carcinoid (<2 mitoses per 2 mm^2^) stage ≥IB with Ki67 <5% and positive CD44 did not develop distant metastases either. All patients with atypical carcinoid (≥2 mitoses 2 mm^2^), Ki-67 of ≥5% (*p*=<0.000) and loss of CD44 (*p*=<0.0001) developed distant metastases. Tumors with stage ≥IB and either ≥2 mitoses, Ki-67 >5% or loss of CD44 metastasized occasionally (11%).

**Conclusion:** Adding tumor diameter, CD44 and Ki-67 to the widely used TC/AC classification, provides a multimodal biomarker that better stratifies patients in prognostically favorable and unfavorable categories than current standards. These findings enable risk stratification allowing a tailored treatment approach for patients with bronchial carcinoid.

## Introduction

Pulmonary carcinoid tumors are a subset of neuroendocrine tumors (NETs) and represent 1-2% of all primary lung malignancies with an incidence of 0.2-2 per 100.000 per year (1). Bronchial carcinoid tumors are predominantly intraluminal, slow growing tumors. Surgical resection is the gold standard for treatment (2). A drawback of anatomical resection in centrally located tumors is the extensive loss of lung parenchyma. Endobronchial therapy (EBT) has emerged as a safe and effective minimally invasive tissue sparing alternative for small intraluminal tumors (3, 4). To date, no randomized controlled trials have compared EBT with surgery for small bronchial carcinoids. This lack of evidence understandably impedes the implementation of EBT. However, an adequately powered RCT would be time consuming because of the low incidence and the low expected number of events (loco regional or distant recurrences). In addition, the indolent growth of carcinoid tumors requires at least 10 years of follow-up. Accurate surrogate biomarkers may be of great help in this setting. Identification of patients with low-risk carcinoids, that do not disseminate, will support the selection of patients suitable for local treatment (surgery or EBT). In addition, lifelong follow-up after local treatment may be unnecessary for such low-risk patients. Conversely, identification of subjects at risk for dissemination may result in more aggressive treatment and/or follow-up strategy.

The best known biomarker in carcinoid is the mitotic index, which is part of the current WHO classification (5). The distinction between typical (TC) and atypical carcinoid (AC) is based on mitotic count and dot-like necrosis in the tumor (1). A tumor with 0-2 mitosis per 2 mm^2^ will be classified as TC and ≥2 mitosis per 2 mm^2^ or necrosis as AC. AC has poorer prognosis and a higher tendency to disseminate (6). Although lymph node involvement and distant metastases predominantly occur in AC, dissemination can also occur in TC (7). In addition, histologic classification in TC and AC is imprecise on small biopsies and should be interpreted with caution (8). Evidently, better biomarkers are required in order to predict prognosis and optimize treatment. Recent studies found additional immunohistochemical markers such as orthopedia homeobox (OTP), CD44 and Ki-67 to have prognostic value in bronchial carcinoid tumors (9) (10, 11) (12, 13). Two other immunohistochemical markers are potential predictors in high grade neuroendocrine carcinoma of the lung, namely retinoblastoma protein (Rb) and P16 (14-16). Most of them are transcription factors, which either activate or repress the gene transcription. Inappropriate expression of these transcription factors might lead to unfavorable cell division, cell growth, or cell death (17, 18).

In the current study, we investigated radiological (tumor diameter), morphometric (mitotic index) and immunohistochemical (OTP, CD44, Ki-67, Rb and P16) markers as predictors of dissemination in a large single institution cohort of patients who underwent treatment for bronchial carcinoid, predominantly with curative intent.

## Materials and Methods

### 1.1 Study design and methods

After approval by the Institutional Review Board (Medical Ethics Review Committee of Amsterdam UMC IRB00002991), patients referred to our tertiary referral center between 1991 and 2019 for treatment of bronchial carcinoid with sufficient tissue for additional immunohistochemistry were included. Patients were selected for EBT or surgery based on detailed information provided by chest CT scans and bronchoscopies. Those patients with evident and significant extraluminal growth, lymph node involvement or multifocal/disseminated disease on CT scan were excluded from EBT and referred for surgery. To detect residual disease, control bronchoscopy and CT scan were typically planned 6 weeks after EBT. Residual disease was confirmed with a biopsy and patients with extensive residual disease were treated with surgery. In patients with minimal residual disease, EBT was repeated if deemed feasible. In the absence of residual disease, patients were followed with annual CT scan and bronchoscopy.

#### 2.1.1 Case evaluation and definitive diagnosis

All cases were analyzed and the diagnosis was confirmed by two pathologists (TR & ET) specialized in lung pathology. Twenty cases were evaluated by both pathologists for concordance testing, discordant cases were discussed during microscope evaluation. Biopsies and surgical tumor specimen where digitally measured to define pathologic tumor size (pT).

#### 2.1.2 Mitotic count and presence of necrosis

Mitotic count and necrosis were scored by a specialized pathologic laboratory technician (MB) as performed in daily diagnostic routine of the pathology ward. In short, the whole slide was first explored for mitotic hotspots and the mitotic count was performed in the hotspot as described before (19). Necrosis was scored by two lung pathologists (TR & ET).

#### 2.1.3 Immunohistochemistry

Immunohistochemistry for Ki-67, Rb, P16, OTP and CD44 was performed. Clones and conditions are presented in Supplementary Table 5. Immunohistochemical staining intensity was scored using the H-score (range 0–300). In short, the percentage of cells at each staining intensity level (0, 1+, 2+, and 3+) was scored, and H-score was calculated using the following formula: 1 × (% cells 1+) + 2 × (% cells 2+) + 3 × (% cells 3+) (20). In markers with overall high expression (OTP, CD44, Rb), the staining intensity of <30 was considered to be negative (21). In markers without or low expression (p16), any staining level was scored as positive. The Ki-67 scoring consisted of estimation of percentage of positive cells in a hotspot region after scanning the whole slide. The highest recorded value was taken into account, as described before (22). The staining intensity of the immunohistochemical markers where analyzed between four prognostic groups (curative treated, recurrence of disease after EBT, lymph node involvement and distant metastases).

### 3.1 Statistical analysis

The statistical analyses and calculations were performed with SPSS 26.0 (IBM Corp., Armonk, N.Y., USA). Chi-square and Fisher tests were applied for comparison of qualitative variables. The area under receiver operating characteristics curve (ROC), abbreviated as AUC, was calculated to identify the discriminatory ability of immunohistochemical markers and mitotic index for recurrence of disease after EBT, lymph node involvement and distant metastases. The Pearson Correlation was used to measure the relationships between pairs of continuous variables (mitotic index and proliferation index). A binary regression analysis was performed to identify independent risk factors for distant metastases. Survival analysis was performed using the Kaplan–Meier method. A *p*-value <0.05 denoted statistical significance.

## Results

### 3.1 patient characteristics

A total of 171 patients were included in the present study. The median age was 49 (interquartile range [IQR] 36-61) and 97 (56.7%) patients were female. TC was diagnosed in 112 (65.5%) and AC in 59 (34.5%) patients (differences in biomarker expression between TC and AC are attached in supplementary Table 3 & 4). A total of 164 (95.9%) patients underwent local treatment with curative intent with either definitive EBT (*n*=61, 35.7%) or surgery (*n*=103, 60.2%). Only 9 patients treated with EBT had a local recurrence of disease. Three patients (33.3%) were treated with EBT (2 patients refused surgery and 1 recurrence was curatively treated with EBT) and 6 (66.6%) were referred for surgical resection. No patients treated with EBT developed distant metastases (follow-up time 75 months (IQR 43-120). A total of 37 (35.9%) of all 103 surgically resected patients (median age 46, IQR 35-59 years) were directly referred for surgery. The remaining 66 (64.1%) patients were initially treated with EBT, but needed surgical resection. Presence of lymph node involvement was detected in 13 patients (12.6%). Also, 13 patients (12.6%) developed distant metastases and most of these metastasized tumors were proven to be AC (11/13, 84.6%). Seven patients (4.1%) presented with distant metastases at diagnosis (liver *n*=2, pleura *n*=2, dura mater *n*=1, skin *n*=1 and adrenal gland *n*=1) (Table 1).

**Table 1:**
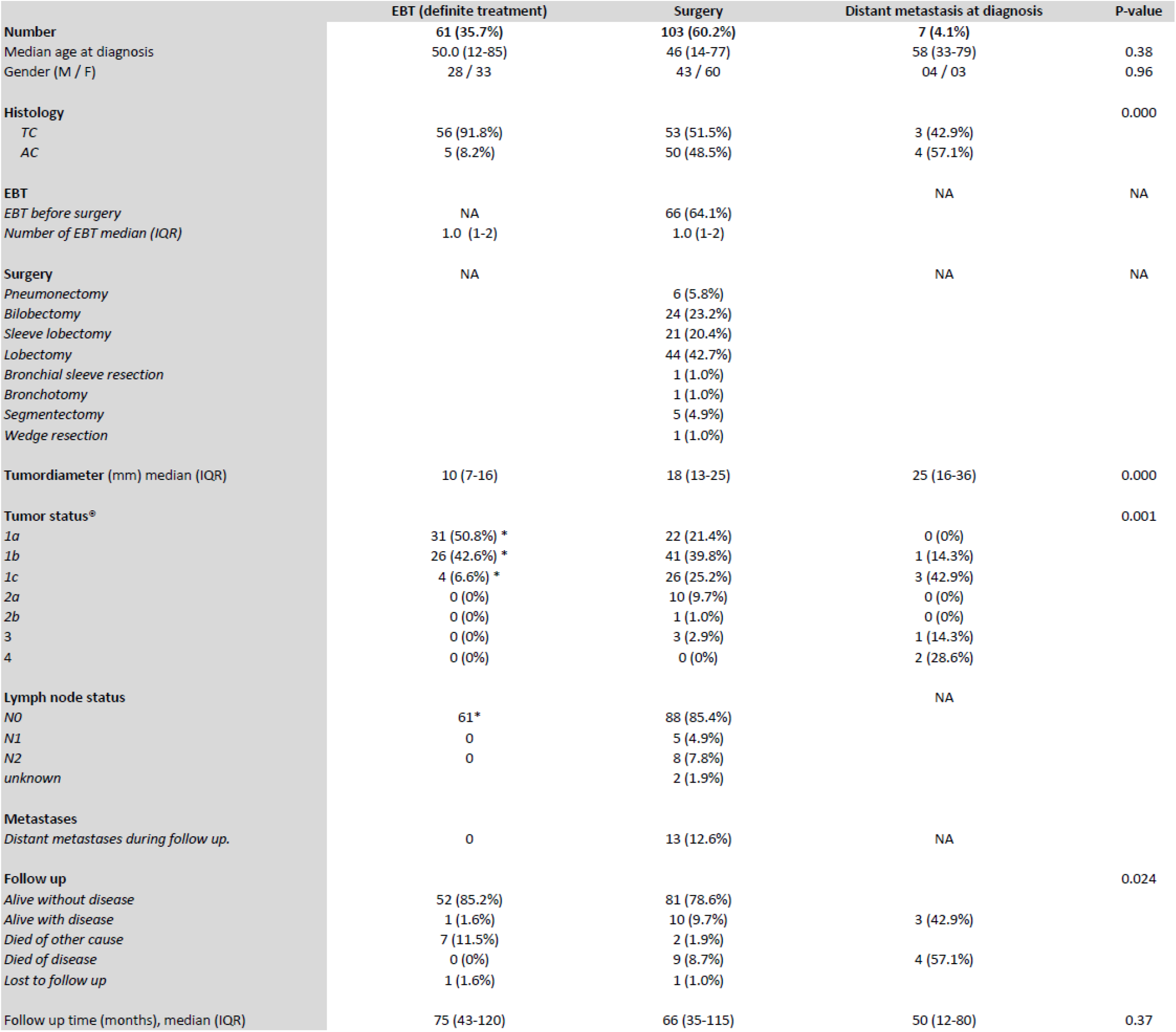
demographics of patients treated for bronchial carcinoid (n=171), ^®^TNM-classification 8^th^ edition. *T and N status based on tumor characteristics on CT scan.

|  | EBT (definite treatment) | Surgery | Distant metastasis at diagnosis | P-value |
| --- | --- | --- | --- | --- |
| **Number** | **61 (35.7%)** | **103 (60.2%)** | **7 (4.1%)** |  |
| Median age at diagnosis | 50.0 (12-85) | 46 (14-77) | 58 (33-79) | 0.38 |
| Gender (M / F) | 28 / 33 | 43 / 60 | 04 / 03 | 0.96 |
| **Histology** |  |  |  | 0.000 |
| TC | 56 (91.8%) | 53 (51.5%) | 3 (42.9%) |  |
| AC | 5 (8.2%) | 50 (48.5%) | 4 (57.1%) |  |
| **EBT** |  |  | NA | NA |
| EBT before surgery | NA | 66 (64.1%) |  |  |
| Number of EBT median (IQR) | 1.0 (1-2) | 1.0 (1-2) |  |  |
| **Surgery** | NA |  | NA | NA |
| Pneumonectomy |  | 6 (5.8%) |  |  |
| Bilobectomy |  | 24 (23.2%) |  |  |
| Sleeve lobectomy |  | 21 (20.4%) |  |  |
| Lobectomy |  | 44 (42.7%) |  |  |
| Bronchial sleeve resection |  | 1 (1.0%) |  |  |
| Bronchotomy |  | 1 (1.0%) |  |  |
| Segmentectomy |  | 5 (4.9%) |  |  |
| Wedge resection |  | 1 (1.0%) |  |  |
| **Tumordiameter (mm) median (IQR)** | **10 (7-16)** | **18 (13-25)** | **25 (16-36)** | **0.000** |
| **Tumor status<sup>a</sup>** |  |  |  | **0.001** |
| 1a | 31 (50.8%) * | 22 (21.4%) | 0 (0%) |  |
| 1b | 26 (42.6%) * | 41 (39.8%) | 1 (14.3%) |  |
| 1c | 4 (6.6%) * | 26 (25.2%) | 3 (42.9%) |  |
| 2a | 0 (0%) | 10 (9.7%) | 0 (0%) |  |
| 2b | 0 (0%) | 1 (1.0%) | 0 (0%) |  |
| 3 | 0 (0%) | 3 (2.9%) | 1 (14.3%) |  |
| 4 | 0 (0%) | 0 (0%) | 2 (28.6%) |  |
| **Lymph node status** |  |  | NA |  |
| N0 | 61* | 88 (85.4%) |  |  |
| N1 | 0 | 5 (4.9%) |  |  |
| N2 | 0 | 8 (7.8%) |  |  |
| unknown |  | 2 (1.9%) |  |  |
| **Metastases** |  |  |  |  |
| Distant metastases during follow up. | 0 | 13 (12.6%) | NA |  |
| **Follow up** |  |  |  | 0.024 |
| Alive without disease | 52 (85.2%) | 81 (78.6%) |  |  |
| Alive with disease | 1 (1.6%) | 10 (9.7%) | 3 (42.9%) |  |
| Died of other cause | 7 (11.5%) | 2 (1.9%) |  |  |
| Died of disease | 0 (0%) | 9 (8.7%) | 4 (57.1%) |  |
| Lost to follow up | 1 (1.6%) | 1 (1.0%) |  |  |
| Follow up time (months), median (IQR) | 75 (43-120) | 66 (35-115) | 50 (12-80) | 0.37 |

### 3.2 Mitotic index

Diagnostic material of patients with distant metastases showed an increased mitotic rate (mean 5.5, standard deviation (SD) ± 5.3) compared to the samples of curatively treated patients (1.2; SD ± 1.7). A mitotic count of at least 2 per 2 mm^2^ (*p*=0.000) was associated with development of distant metastases (AUC 0.76, *p*=0.002) (Figure 2A).

**Figure 1:**
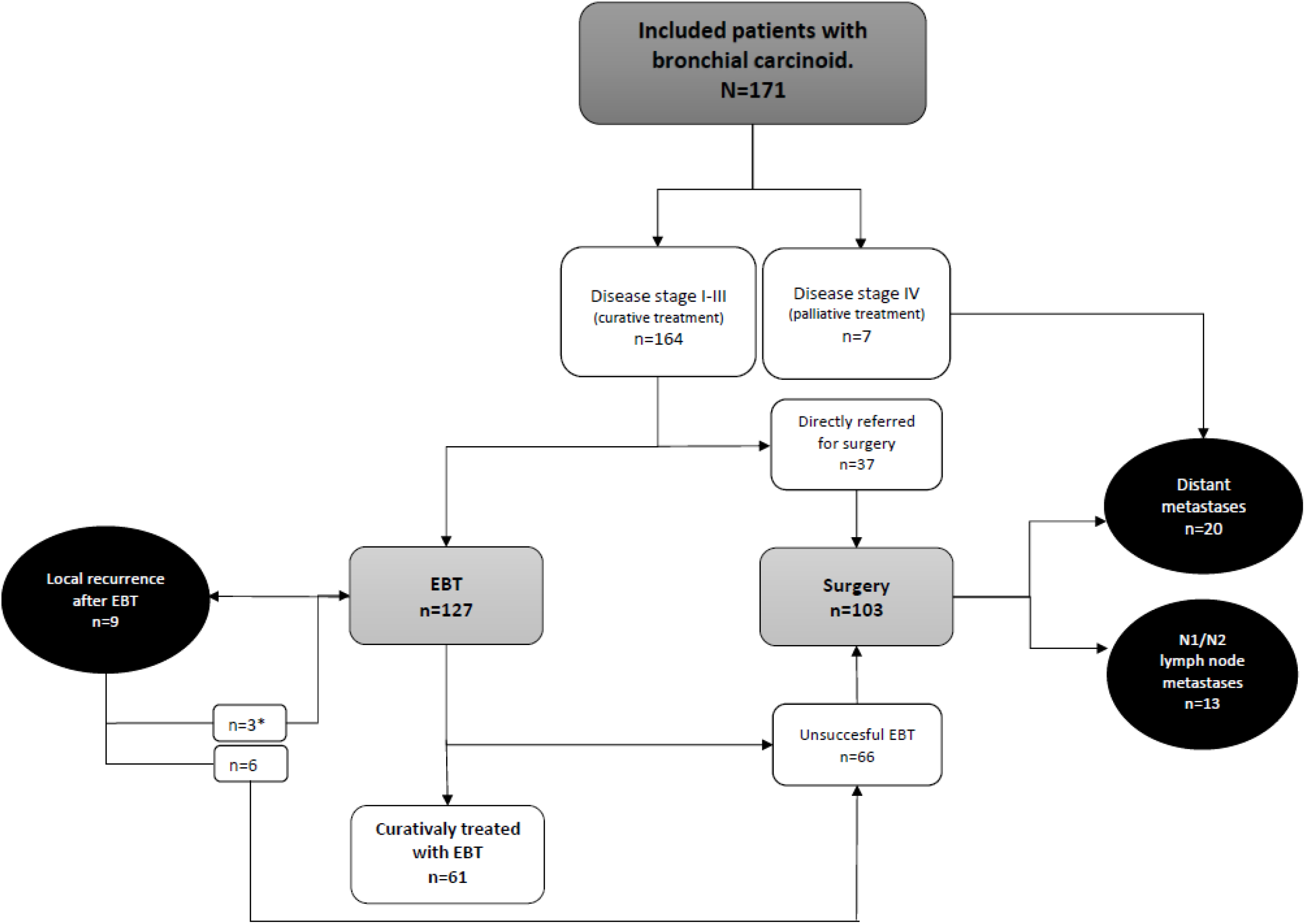
flowchart of 171 patients treated with bronchial carcinoid, * two patients refused surgery and 1 recurrence was curatively treated with EBT.

**Figure 2:**
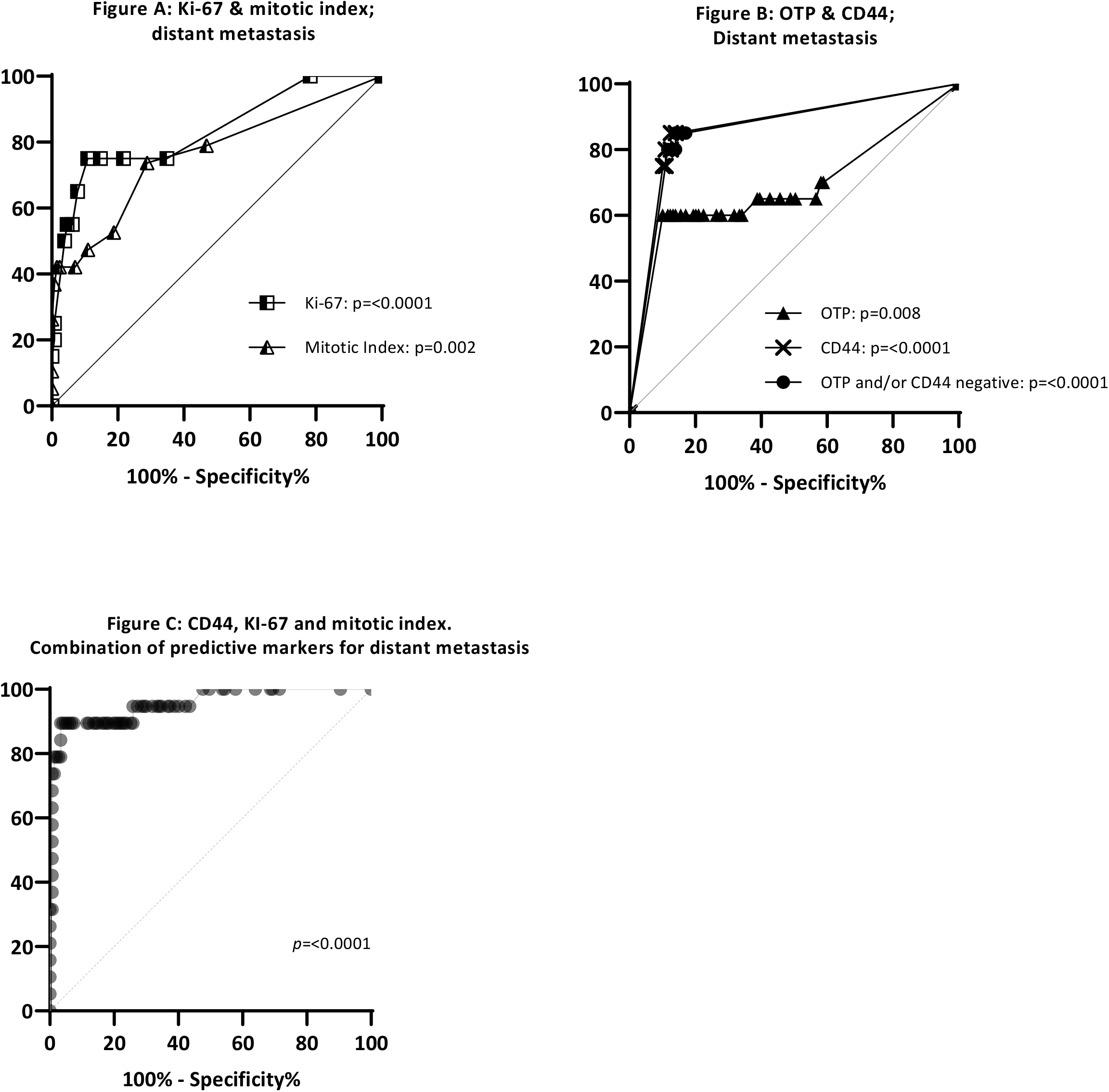
Ki-67 (AUC 0.83) and mitotic index (AUC 0.76) in patients with distant metastases (n=20) compared to patient without distant metastases (n=129) (Figure A). OTP (AUC 0.69), CD44 (AUC 0.87) of OTP and/or CD44 negative (AUC 0.86) in patient with distant metastases (n=20) compared to patient without distant metastases (n=129) (Figure B). Combination of predictive markers; negative CD44, Ki-67 and mitotic index (>2 mm^2^) for distant metastases (AUC 0.96) (Figure C).

### 3.3 Ki-67

The mean Ki-67 proliferation index of all cases in this cohort was 2.8% (SD ± 3.8). Ki-67 tended to be higher in AC than in TC (4% (SD ± 5.0) vs 2% (SD ± 3.0), *p*=0.06) (supplementary Table 3). Mitotic count and Ki-67 showed no correlation (r=0.1, p=0.22) (Supplementary Figure 4). A relatively high Ki-67 index was observed in patients with N1 and N2 lymph node involvement (3.2%; SD ± 3.0) and distant metastases (8.6%; SD ± 6.4) compared to curative treated patients (1.9%; SD ± 2.5). The ROC analysis confirmed that an increased Ki-67 is discriminant for developing distant metastases (AUC 0.83; *p*<0.0001) (Figure 2A). A Ki-67 threshold of ≥5% (*p*<0.000) was also associated with increased risk of distant metastases.

### 3.4 Rb and P16

Rb was positive in 90% of the patients. There was no significant difference for Rb between the 4 prognostic groups (curative treated, recurrence of disease after EBT, lymph node involvement and distant metastases). P16, largely inversely associated to Rb, was positive in 16% of all patients ^13^. Comparing the 4 prognostic groups, patients with distant metastases showed p16 positivity more often (30% of the cases). However, this did not reach statistical significance (*p*=0.18) (Table 2).

**Table 2:** number of positive staining, area under the curve (AUC) of the ROC curves and P-values of different immunohistochemical staining in four prognostic groups. Marked P-value are significant values.*CD44 (n=2) and P16 (n=1) missing.

| Marker | Number of positive tumors | % | AUC | P-value |
| --- | --- | --- | --- | --- |
| **Rb** |  |  |  |  |
| **Whole group** | **154/171** | **90%** | **NA** | **NA** |
| Curative treatment (EBT, surgery, N0) | 116/129 | 90% | NA | NA |
| Local recurrence of disease after EBT | 8/9 | 89% | 0.57 | 0.48 |
| N1 or N2 lymph node metastases | 13/13 | 100% | 0.66 | 0.60 |
| Distant metastases | 17/20 | 85% | 0.52 | 0.79 |
| **P16** |  |  |  |  |
| **Whole group** | **27/170\*** | **16%** | **NA** | **NA** |
| Curative treatment (EBT, surgery, N0) | 15/129 | 12% | NA | NA |
| Local recurrence of disease after EBT | 4/9 | 11% | 0.65 | 0.13 |
| N1 or N2 lymph node metastases | 2/12* | 17% | 0.45 | 0.60 |
| Distant metastases | 6/20 | 30% | 0.59 | 0.18 |
| **OTP** |  |  |  |  |
| **Whole group** | **141/171** | **83%** | **NA** | **NA** |
| Curative treatment (EBT, surgery, N0) | 114/129 | 88% | NA | NA |
| Local recurrence of disease after EBT | 7/9 | 77% | 0.54 | 0.68 |
| N1 or N2 lymph node metastases | 12/13 | 92% | 0.60 | 0.85 |
| Distant metastases | 8/20 | 40% | 0.69 | 0.008 |
| **CD44** |  |  |  |  |
| **Whole group** | **139/169\*** | **82%** | **NA** | **NA** |
| Curative treatment (EBT, surgery, N0) | 115/128* | 90% | NA | NA |
| Local recurrence of disease after EBT | 7/8* | 77% | 0.51 | 0.95 |
| N1 or N2 lymph node metastases | 12/13 | 92% | 0.41 | 0.07 |
| Distant metastases | 5/20 | 25% | 0.84 | <0.0001 |
| **OTP and/or CD44 negative** |  |  |  |  |
| **Whole group** | **131/169\*** | **78%** | **NA** | **NA** |
| Curative treatment (EBT, surgery, N0) | 110/128* | 86% | NA | NA |
| Local recurrence of disease after EBT | 6/8* | 75% | 0.55 | 0.67 |
| N1 or N2 lymph node metastases | 12/13 | 93% | 0.44 | 0.55 |
| Distant metastases | 3/20 | 15% | 0.85 | <0.0001 |
| Ki-67 |  |  |  |  |
| **Whole group** | **NA** | **NA** | **NA** | **NA** |
| Curative treatment (EBT, surgery, N0) | NA | NA | NA | NA |
| Local recurrence of disease after EBT | NA | NA | 0.54 | 0.68 |
| N1 or N2 lymph node metastases | NA | NA | 0.60 | 0.22 |
| Distant metastases | NA | NA | 0.83 | <0.0001 |
| Mitotic index |  |  |  |  |
| **Whole group** | **NA** | **NA** | **NA** | **NA** |
| Curative treatment (EBT, surgery, N0) | NA | NA | NA | NA |
| Local recurrence of disease after EBT | NA | NA | 0.55 | 0.60 |
| N1 or N2 lymph node metastases | NA | NA | 0.31 | 0.29 |
| Distant metastases | NA | NA | 0.76 | 0.002 |

### 3.5 OTP and CD44

In the whole cohort loss of OTP and CD44 expression was found in 17% and 18%, respectively. In patients with distant metastases the loss of expression of OTP and CD44 was significantly higher (OTP; 60%; *p*=0.008, CD44; 75%, *p*=0.0001). In patients with N1 or N2 lymph node involvement or recurrence of disease after EBT no significant loss of OTP or CD44 expression was observed. ROC analysis revealed a slightly higher area under the curve (AUC) value for CD44 (0.87) than for OTP (0.68) Combining the OTP and CD44 data did not increase the ability to discriminate between patients with and without distant metastases (AUC: 0.86), (Figure 2B).

### 3.6 Risk factors distant metastases

Subsequently, binary logistic regression was employed to identify a multivariate panel of risk factors of distant metastases in patients with bronchial carcinoid. The results of this analysis revealed that loss of CD44 (*p*=<0.000), immunohistochemistry for Ki-67 (*p*=0.002) and mitotic count (*p*=0.018) were independently associated with a higher risk of distant metastases. In comparison to individual markers, this combination of predictors demonstrated the greatest performance in terms of discriminatory capacity (*p*=0.0001, AUC 0.96) (Figure 2D).

### 3.7 Survival

Finally, a survival analysis was performed to assess the influence of pathologic tumor diameter (pT) on the development of distant metastases. In the pT stage IA bronchial carcinoid tumors the disease free survival rate was 100% (Figure 3A). Within the patients with pT stage IB-IV, histology of AC (≥2 mitosis per 2 mm^2^), loss of CD44 and Ki-67 ≥5% showed prognostic value for development of distant metastases (Figure 3B, p= <0.0001). If all three factors were present the 50% disease free survival rate was 48 months. If one of these unfavorable prognostic characteristics was present 11% (6/54 patients) metastasized, one patient with TC and five AC. This patient group is further specified in (supplementary Table 4).

**Figure 3:**
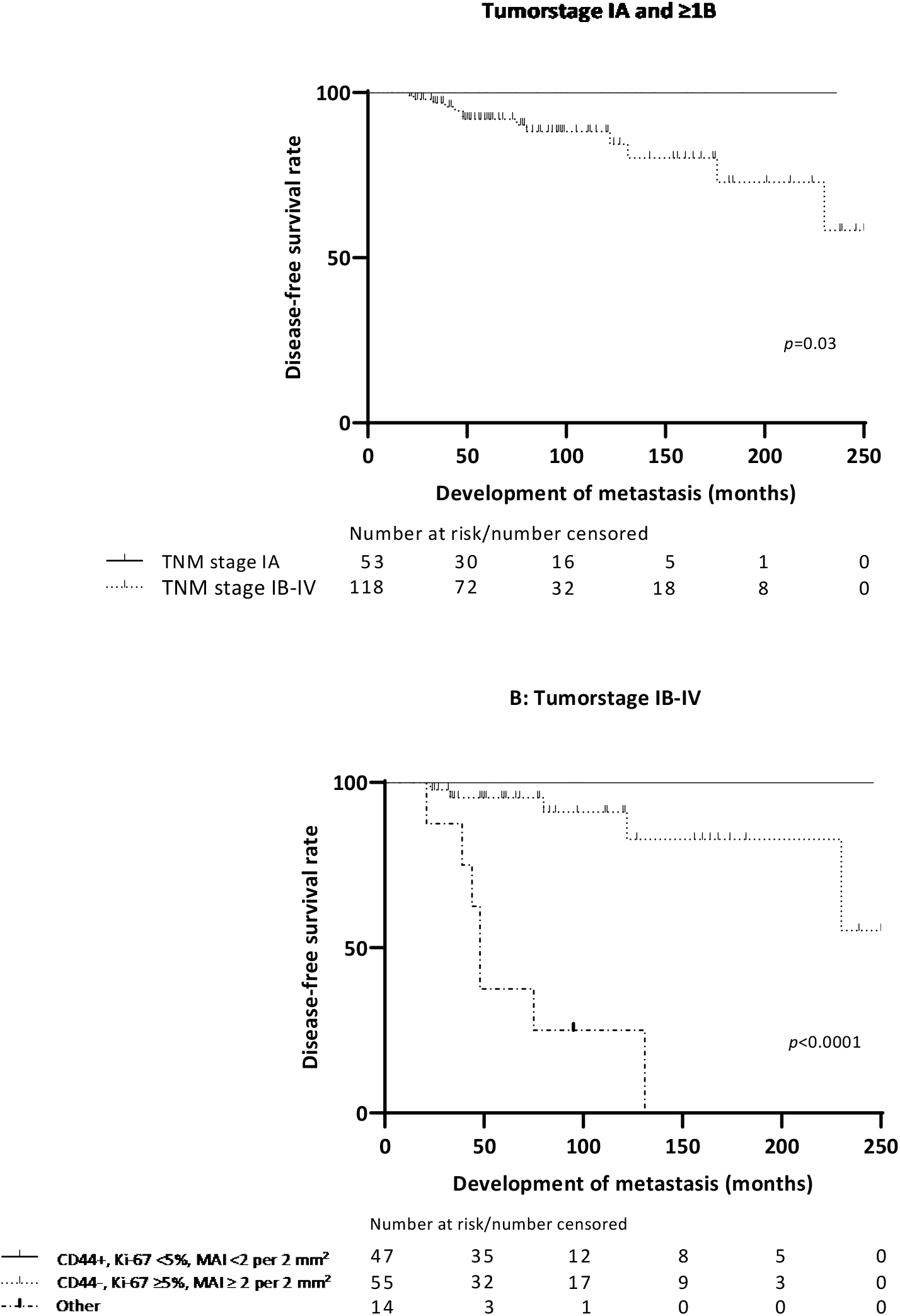
Survival curves. Development of distant metastases based on pathologic tumorstage (pT) for all patients divided in pT IA and IB-IV (A) and for pT ≥IB based on the combination of the 3 prognostic immunohistochemical markers (two patients were missing in CD44) (B).

## Discussion

In this study we demonstrate that combining tumor diameter (pT), mitotic count, CD44 and Ki-67 expression greatly improves prediction in patients with bronchial carcinoid. All bronchial carcinoids ≤1 cm and all bronchial carcinoids >1 cm with a low mitotic index (<2 per 2 mm^2^), a low Ki-67 expression (<5%) and a normal CD44 do not disseminate.

This study demonstrated a similar prognostic value for loss of OTP and/or CD44 for developing metastatic disease as previously reported in the literature. Loss of OTP and/or CD44 was independently associated with poor survival and increased risk of metastases (9, 23, 24). Most interestingly, studies showed that OTP in combination with CD44, allowed even better separation of tumors into prognostic relevant categories (23, 25). However, this study showed that CD44 has an identical prognostic value for development of distant metastases in patients with bronchial carcinoid compared to the combination of OTP and CD44. In other words, CD44 alone is a very reliable marker, what has, in contrast to OTP, the advantage of wide availability.

Ki-67 is a nuclear protein which plays an essential role in the control and timing of cell proliferation (12). The prognostic model of our study showed that a Ki-67 of >5% was associated with risk for development of distant metastases, what is in line with a two other studies reporting that a Ki-67 cutoff of 2.5% to 5% differentiates between lower and higher aggressive NE tumors in the setting of TC and AC (10, 12, 26). Remarkably, Ki67 and mitotic count showed no correlation. A possible explanation for this is that Ki-67 protein is present during all active phases of the cell cycle (G1, S, G2, and mitosis), but is absent in resting (quiescent) cells (G0). The mitotic count is only based on the mitotic phase of the cell cycle (27). This supports the additive prognostic value of the two. Ki-67 never exceeded 20% in this carcinoid cohort, a suggested cut-off value for the differential diagnosis with LCNEC (2). Ki-67, together with the mitotic count, is already used in a grading system for gastrointestinal neuroendocrine tumors to better predict the biological behavior of these entities (28).

From our data, we learn that Rb and p16 are not related to the development of metastases or differentiation between TC and AC. Few studies demonstrated Rb function essentially intact in TC’s but lost in approximately 50% of ACs (15, 29). Our data show a rather random loss of Rb in ca 10% of the cases, irrespective of the prognostic group. RB is a low abundant nuclear transcription factor. Lost immunoreactivity might also be a false negative immunohistochemical result, what can be attributed to preanalytical variables (30). Although p16 was partially positive in several cases with distant metastases, it was never diffusely positive. Hypothetically, focal p16 staining in carcinoids may be used as diagnostic marker for the differential diagnosis with LCNEC, where it was often diffusely strong positive (16, 29).

To place our results in clinical context, we should explore how they might change the management of patients with bronchial carcinoid. In order to perform EBT as a minimally invasive treatment in patients with bronchial carcinoid, one would preferably only select patients without a chance of lymph node involvement and without a tendency to develop distant metastases. In the current study, all pT1AN0 tumors did not disseminate, irrespective of the mitotic count and CD44 and Ki67 findings. Also, pT1B tumors with a mitotic count < 2 per 2 mm^2^, a low Ki-67 expression (<5%) and a normal CD44 did not disseminate. These 2 categories of patients may be good candidates for EBT, provided there is no extraluminal component. Consequently, all other patients with resectable disease should then be treated with radical surgical resection. Of course, prospective trials would be required in order to assess whether such a surrogate biomarker-driven treatment strategy is effective and safe.

With regard to follow-up procedure after radical treatment, this biomarker set could also change our strategy. Because local recurrence occurs in a small proportion of patients after EBT, bronchoscopic follow-up is obviously mandatory. But after radical surgical resection of a tumor with a favourable biomarker profile (low mitotic count, low Ki-67 expression and normal CD44 expression), yearly follow-up may be futile.

The findings of the present study must be interpreted in the context of several potential limitations. Firstly, since the patients were referred to our hospital for treatment with EBT, the population included in this study has been exposed to selection bias. Patients treated with EBT were predominantly small TC tumors compared to the group that underwent surgical resection. However, this allowed us to investigate the prognostic value of morphology and immunohistochemistry in the context of low T stage with an intensive and long follow-up. Secondly, as previously stated, ki-67 and mitotic count has interobserver variability (31). Our study is a single-center study, with experienced laboratorian technician (32) and pathologists specialized in mitotic count in a tertiary referral center. Therefore, these results should be validated in other patient groups outside our institution. Another limitation is the median follow-up time of 5 years, considering the indolent growth of these tumors. Long term results should be analyzed in the future.

## Conclusion

Adding tumor diameter, CD44 and Ki-67 to the widely used TC/AC classification, provides us with a multimodal biomarker that seems to accurately stratify patients in favorable and unfavorable groups. pT1AN0 tumors, irrespective of MAI or immunohistochemistry findings and pT1B tumors with a favorable biomarker profile (MAI <2 per 2 mm^2^, Ki-67 expression (<5%) and a positive CD44) did not metastasize. In the future, these findings may enable risk stratification in order to provide a tailored approach with regards to treatment and follow-up of patients with bronchial carcinoid.

## Data Availability

The data that support the findings of this study are available from the corresponding author, [EMBP Reuling], upon reasonable request. However, there might be restrictions e.g. containing information that could compromise the privacy of research

## Acknowledgements

ER, TR, ET, and HD were responsible for the conception and design of the study and acquisition of data. ER ET, and DN performed analysis and interpretation of the data. The article has been written by ER, TR, DN critically revised by the authors who all gave approval for submission. Each author has participated sufficiently in the contributions of this article and agreed to be accountable for all aspects of the work in ensuring that questions related to the accuracy or integrity of any part of the work are appropriately investigated and resolved. This study was supported by a grant of ORAS (Oncological Research Albert Schweitzer Hospital). We thank Kartik Viswanathan for sharing OTP antibody from Weill Cornell Medicine, Department of Pathology and Laboratory medicine.

