## Supplemental Table 3-4-5 and Supplemental Figure 4 for "A multimodal biomarker predicts dissemination of bronchial carcinoid"

***Supplemental data article: "A multimodal biomarker predicts dissemination of bronchial carcinoid". EMBP Reuling et al.***

| | **TC** (n/total) | **%** | **AC** (n/total) | **%** | **Total** (n/total) | **%** | **P-value** |
|---|---|---|---|---|---|---|---|
| **Rb +** | 96/111 | 86.5% | 56/59 | 94.9% | 152/170 | 89.4% | 0.09 |
| **P16 +** | 30/111 | 27.0% | 22/59 | 37.3% | 52/170 | 30.6% | 0.17 |
| **OTP +** | 95/112 | 84.8% | 45/59 | 76.3% | 140/171 | 81.9% | 0.17 |
| **CD44 +** | 97/111 | 87.4% | 40/58 | 70.0% | 137/169 | 81.1% | 0.004 |
| **Ki-67 (mean/SD)** | 2 (3.0) | NA | 4 (5.0) | NA | 3 (3.8) | NA | 0.06 |

*Supplementary Table 3: amount of positive stainings between TC and AC.*

| | **Totaal (n=55)** | **TC (n=17)** | **AC (n=38)** | ***p*-value** |
|---|---|---|---|---|
| **CD44 postive** | 42 | 10 | 32 | 0.08 |
| **CD44 negative** | 13 | 7 | 6 | |
| **Ki-67 <5%** | 43 | 8 | 35 | 0.000 |
| **Ki-67 ≥5%** | 12 | 9 | 3 | |
| **Metastases** | 6 | 1 | 5 | 0.65 |
| **No metastases** | 49 | 16 | 33 | |

*Supplementary Table 4: Patients (n=55) with one of the prognostic unfavorable characteristics ("other-group") (negative CD44, Ki-67) in relation to TC and AC.*

| **Antibody** | **Company name** | **Clone** | **Species** | **Reference number** | **Dilution** | **IHC Incubation time (min)** | **Detection method** |
|---|---|---|---|---|---|---|---|
| **CD044** | Ventana | SP37 | Rb | 06364985001 | RTU | 32 | Optiview |
| **MIB 1** | Dako | MIB1 | Mo | M7240 | 1/50 | 32 | Optiview |
| **OTP** | Sigma | polycl | Rb | HPA059342 | 1/50 | 64 | Optiview |
| **RB** | BD biosciences | G3-245 | Mo | 554136 | 1/1000 in Dako reduc ab dil | 32 | Optiview |
| **P16** | Ventana | E6H4 | Mo | 06695248001 | 1/2 | 32 | Optiview |

*Supplementary Table 5: Clones and conditions of immunohistochemical markers, IHC: immunohistochemistry.*

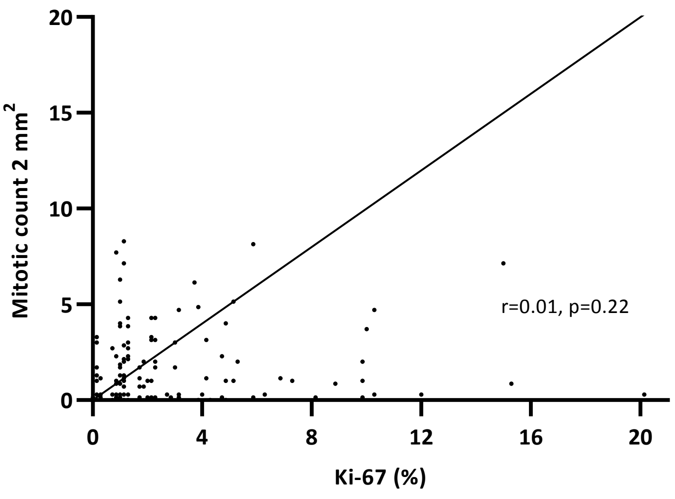

*Supplementary Figure 4: correlation between Ki-67 and mitotic count, p=0.22.*
